# Co-occurrence patterns and risk factors of migraine and attention deficit hyperactivity disorder across 204 countries and territories: a systematic analysis of the Global Burden of Disease Study

**DOI:** 10.64898/2026.05.13.26352990

**Authors:** Xianbing Wang, Jiaying Jiang

## Abstract

**Background:** Migraine prevalence is higher among individuals with attention deficit hyperactivity disorder (ADHD). However, most research has focused on single-disease studies. This study used Global Burden of Disease (GBD) data to analyze co-occurrence patterns and related risk factors.

**Methods:** This study extracted the incidence and age-standardized incidence rate (ASIR) of migraine and ADHD among individuals across 204 countries and territories in GBD 2021, as well as exposure values for risk factors. To explore the co-occurrence patterns of migraine and ADHD and their spatial heterogeneity in global distribution, the incidence of both diseases was classified into quartiles, and countries and territories were categorized into three regional types: consistent regions, migraine-dominant regions, and ADHD-dominant regions. Global groupings by economy and risk factors were analyzed separately for co-occurrence patterns, and disease burden projections were made for 2050.

**Results:** In 2021, countries and regions were categorized into three distinct groups based on disease prevalence patterns: the majority exhibited an ADHD-dominant profile, predominantly found in high-SDI regions; a consistent pattern, where both diseases occurred at comparable levels, was primarily observed across South Africa and the Middle East, while a migraine-dominant pattern was identified in North Africa. Co-occurrence patterns were generally less prevalent in areas with lower socioeconomic development. Across all three patterns, high temperature exposure, iron deficiency, and metabolic risks emerged as the primary contributing factors. Looking ahead to 2050, the global burden of migraine was projected to stabilize, whereas the prevalence of ADHD was expected to experience a slight yet consistent increase.

**Conclusion:** This study systematically identifies the co-occurrence patterns of ADHD and migraine, along with their socioeconomic and environmental drivers, offering evidence-based insights for early prevention and targeted intervention in disease populations globally.

## 1 Introduction

Attention-deficit/hyperactivity disorder (ADHD) and migraine are both neurological disorders associated with substantial disease burdens^[1][2]^. Childhood is the most common period for ADHD diagnosis particularly the age range of 5–9 years, and migraine also shows a relatively high occurrence during this developmental stage^[3][4]^. Data from the Global Burden of Disease (GBD) study indicate that, from 1990 to 2021, the global incidence, prevalence, and disability-adjusted life years (DALYs) of ADHD increased by 9.92%, 18.71%, and 18.57%, respectively^[5]^. During the same period, the global burden of migraine also increased markedly, with the number of cases rising from 732.56 million to 1.16 billion, the prevalence rate increasing by 42.06%, and DALYs increasing by 58.27%^[6]^. Against this growing epidemiological background, the economic burden caused by these two disorders is considerable^[6][7]^.

Notably, multiple meta-analyses and epidemiological studies have suggested a positive association between ADHD and migraine^[8][9][10]^. In addition, the two disorders share several comorbidity patterns, including anxiety and depression, which suggests that common pathophysiological mechanisms may underlie their co-occurrence^[11]^. However, the global temporal trends, spatial distribution, and modifiable risk factors associated with the comorbidity patterns of ADHD and migraine remain insufficiently and unsystematically evaluated.

The GBD study aims to quantify the impact of diseases, injuries, and risk factors on global health^[12]^. Therefore, using the latest data from GBD 2021, this study evaluates the global disease burden trends of comorbid ADHD and migraine over the past three decades. By focusing on comorbidity and modifiable risks, this study aims to characterize the epidemiological evolution of these conditions and to provide comprehensive evidence and predictive insights for optimizing clinical prevention and management strategies and reducing the overall disease burden.

## 2 Materials and methodology

### 2.1 Data acquisition

Data on the incidence of migraine and attention deficit hyperactivity disorder (ADHD) were obtained from the Global Burden of Diseases (GBD) 2021. In brief, the GBD systematically integrates global data on 369 diseases and injuries and 88 risk factors. It provides comprehensive assessments across age groups, sexes, geographical regions, and time periods, including estimates of temporal trends, disease burden, and risk attribution. The GBD utilizes the International Classification of Diseases (ICD) system to code and classify diseases, thereby facilitating the interpretation and comparison of global health trends. The Institutional Review Board at the University of Washington approved an exemption from the need for informed permission to access the GBD data, and the Guidelines for Accurate and Transparent Health Estimates Reporting (GATHER) were adhered to in this investigation. This study extracted the incidence, along with the corresponding 95% uncertainty intervals (UIs), of migraine and ADHD among individuals across 204 countries and territories in GBD 2021, as well as exposure values for risk factors at the most detailed level of definition, encompassing environmental, occupational, and behavioral domains. This dataset, which spans the years 1990–2021, includes the age group from 5 to 9 years.

### 2.2 Socio-demographic index (SDI)

The social and economic factors influencing health outcomes in various regions are reflected in the SDI, a composite metric. It is the geometric mean of three indicators: the average education level for people aged 15 and over (EDU15+), the per capita lag-distributed income (LDI), and the total fertility rate (TFR) for people under 25 (TFU25). The 204 countries and territories were categorized into SDI quintiles ^[13]^.

### 2.3 Co-occurrence region division of migraine and ADHD

To explore the co-occurrence patterns of migraine and ADHD and their spatial heterogeneity in global distribution, the incidence of both diseases was classified into quartiles. Based on the incidence levels in same year of migraine and ADHD, countries and territories were categorized into three regional types: consistent regions (where both diseases share the same incidence level), migraine-dominant regions (where migraine incidence is higher), and ADHD-dominant regions (where ADHD incidence is higher). Based on the incidence for migraine and ADHD across countries from the 1990-2021 dataset, a scatter plot of country-year points was generated with migraine incidence on the x-axis and ADHD incidence on the y-axis. LOESS smoothing curves were fitted for each category separately. Spearman’s correlation was used to estimate the correlation coefficient and corresponding *P*-value between the burdens of the two disorders. Based on the incidence of both diseases from the GBD 1990–2021 dataset, a co-occurrence index was calculated for each country-year unit by averaging the incidence of the two diseases, followed by scale normalization. The index was then discretized into four severity levels (1–4) according to its quartiles.

### 2.4 Identification of risk factors for co-occurrence patterns of migraine and ADHD

The regions were grouped according to three co-occurrence patterns. A random forest model was first trained to identify the risk factors with the greatest contribution to the grouping, followed by an interpretability analysis using TreeSHAP to obtain the direction and magnitude of the marginal contribution (SHAP values) of each risk factor to the classification, along with comparisons across groups.

### 2.5 Disease burden prediction

Utilizing the age-standardized incidence rate (ASIR) for children aged 5–9 years from the GBD 1990–2021 dataset, time series were constructed for each country, stratified by sex and disease (migraine and ADHD). Each time series was differenced to achieve stationarity. Optimal ARIMA(p,d,q) orders were selected based on ACF/PACF patterns and minimization of AIC/BIC values. The residuals were tested for white noise using the Ljung–Box test (*P* > 0.05). The fitted models were used to forecast incidence rates from 2022 to 2050, along with 80% and 95% prediction intervals (PI).

### 2.6 Statistical analysis

We examined burden variations overall and in 204 countries and territories, as well as through demographic stratification by gender and development levels (SDI quintiles), with emphasis on cross-sectional data from 1990 and 2021, along with the estimated annual percentage changes (EAPCs) of ASIR from 1990 to 2021. This study employed DisMod-MR 2.1 to estimate incidence. The tool integrates multiple disease parameters, epidemiological associations, and geospatial data within a Bayesian statistical framework to generate highly robust estimation results. The formula for calculating the age-standardized rate (ASR) is as follows:

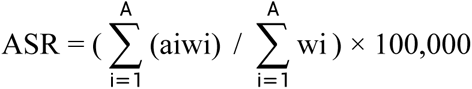

wi: the people number in the matching ith age group among the standard population; ai: the age-specific rate in ith age group;

A: the number of age groups

The EAPC is analyzed based on a regression model used to describe the variation pattern of the ASR over a specific time period. The regression equation applied is as follows: Y = α + βX + e . Here, Y represents the natural logarithm of the ASR, X corresponds to the calendar year, α denotes the intercept, β is the slope (i.e., the trend of change), and e is the random error term. The EAPC is calculated using the formula 100 × [exp(β) – 1], reflecting the annual percentage change^[14]^. All analyses were implemented in R version 4.2.2.

## 3 Results

### 3.1 Global burden of migraine and ADHD

In 2021, the highest incidence of migraine was observed in South America and parts of Europe, while incidence in Africa and Southeast Asia was relatively lower. In contrast, the burden of ADHD was most pronounced in Australia and certain European regions, with consistently lower rates across Africa (**Figure 1A-B**). Between 1990 and 2021, ADHD ASIR showed a mild upward trend globally, with more noticeable increases in East Asia and some European countries. Meanwhile, migraine trends exhibited considerable regional variation, with heterogeneous patterns across countries—some experiencing rising ASIR, others remaining stable, and a number showing declining rates (**Figure 1C-D**).

**Figure 1.**
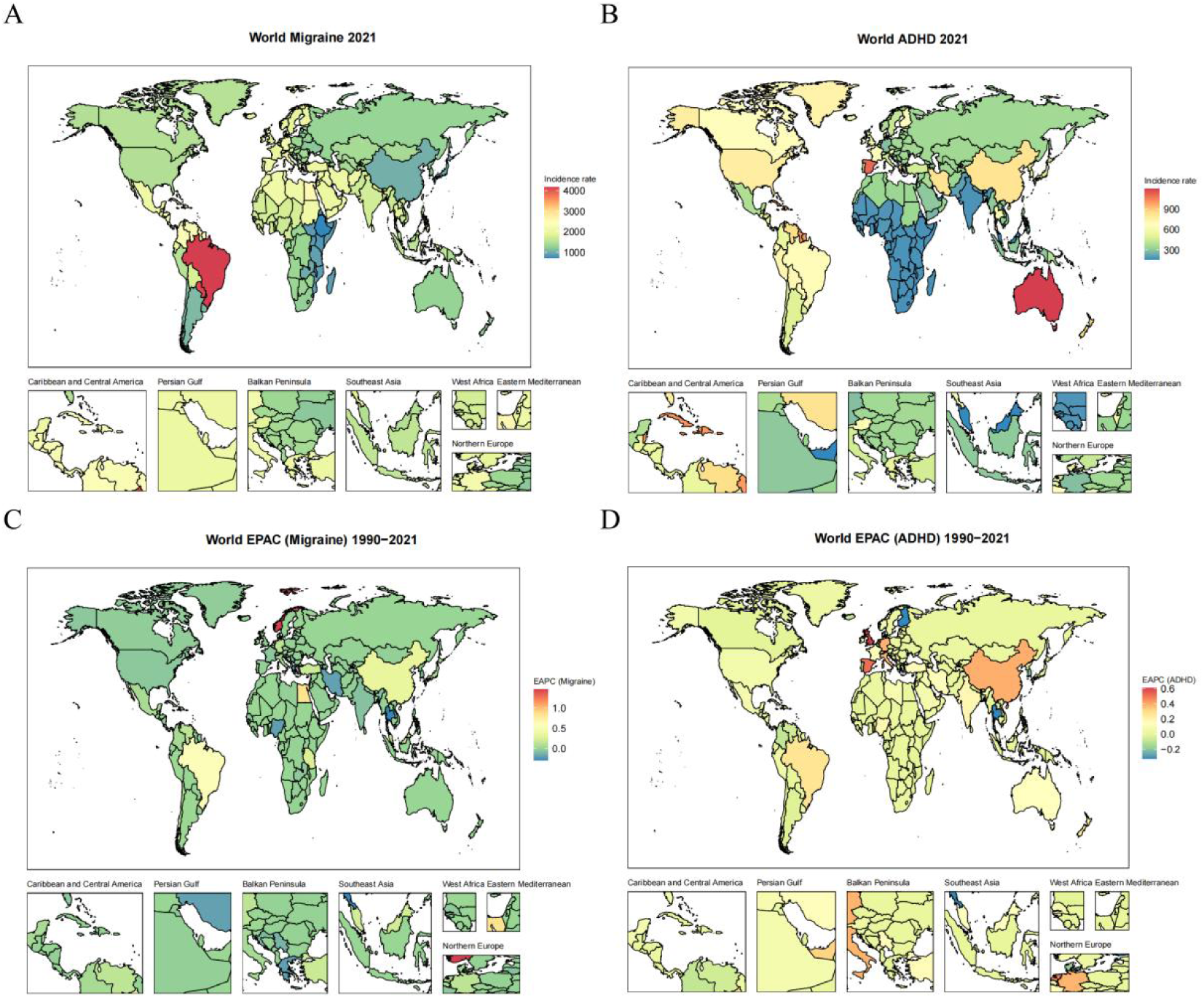
Global burden of migraine and attention deficit hyperactivity disorder (ADHD). (**A-B**) Global distribution of the incidence of migraine (**A**) and ADHD (**B**) in 2021. (**C-D**) Global distribution of estimated annual percentage changes (EAPCs) of in age-standardized incidence rate (ASIR) of migraine (**C**) and ADHD (**D**) from 1990 to 2021.

From 1990 to 2021, the global ASIR of migraine showed a slight increase from 1606.6 to 1612.8 per 100,000 population, with an EAPC of 0.08% (0.03–0.14), indicating a mild upward trend overall (**Table 1**). In terms of absolute burden, the number of incidence globally rose from approximately 9,375,144 in 1990 to 11,080,972 in 2021 per 100,000 population. A clear sex-based disparity was observed. The ASIR for females was markedly higher than that for males in 2021 (2031.0 vs. 1220.4 per 100,000). However, the trend for females was not marked (EAPC = 0.04 (−0.03 to 0.1)), while a notable increase was seen in males (EAPC = 0.16 (0.11–0.21)). When stratified by the SDI, distinct epidemiological patterns emerged: high-middle and middle SDI regions experienced the most substantial increases in incidence (EAPC = 0.30 and 0.18, respectively), while high-SDI regions saw a more modest rise (EAPC = 0.09). In contrast, low-middle SDI regions remained relatively stable (EAPC = 0.00) and low-SDI regions exhibited a slight decline (EAPC = -0.04). Notably, the low-middle SDI region carried the highest disease burden in 2021, with an ASIR of 1830.9 per 100,000. Illustrating these trends, high-income North America—a high-SDI region—reported an ASIR of 1478.87 per 100,000 in 2021 with a slight decrease over the period (EAPC = -0.05%), whereas South Asia, a low-middle SDI region, maintained a high but stable burden (ASIR = 1731.38) with a slight decreasing trend (EAPC = -0.06%). Meanwhile, Western Sub-Saharan Africa, representing low SDI, demonstrated a significant decline (EAPC = -0.09%). This analysis underscores significant disparities by development level, indicating a concentration of increasing burden in middle and high-middle SDI regions, while certain low and low-middle SDI regions show signs of stabilization or decline.

**Table 1.**
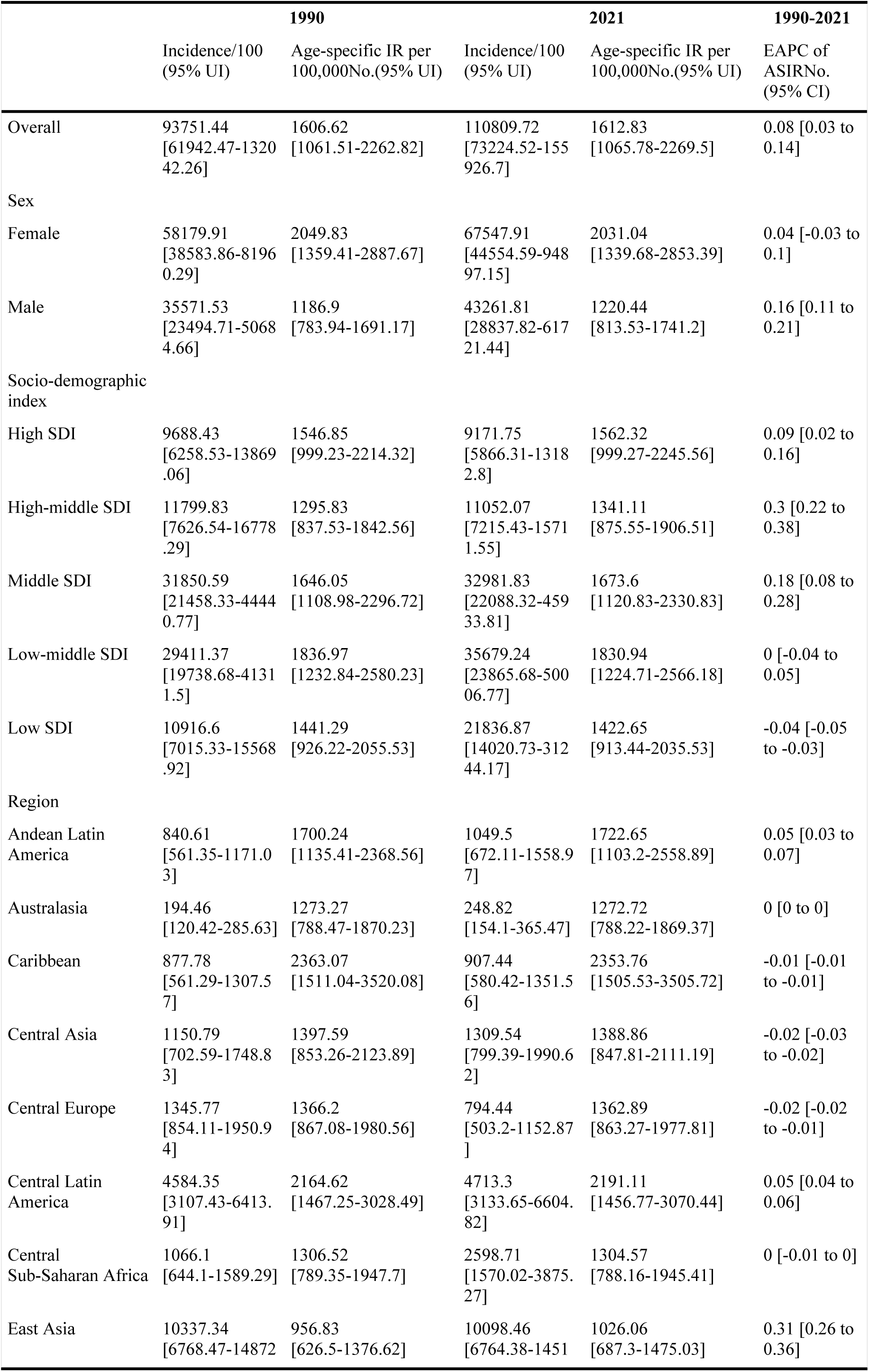
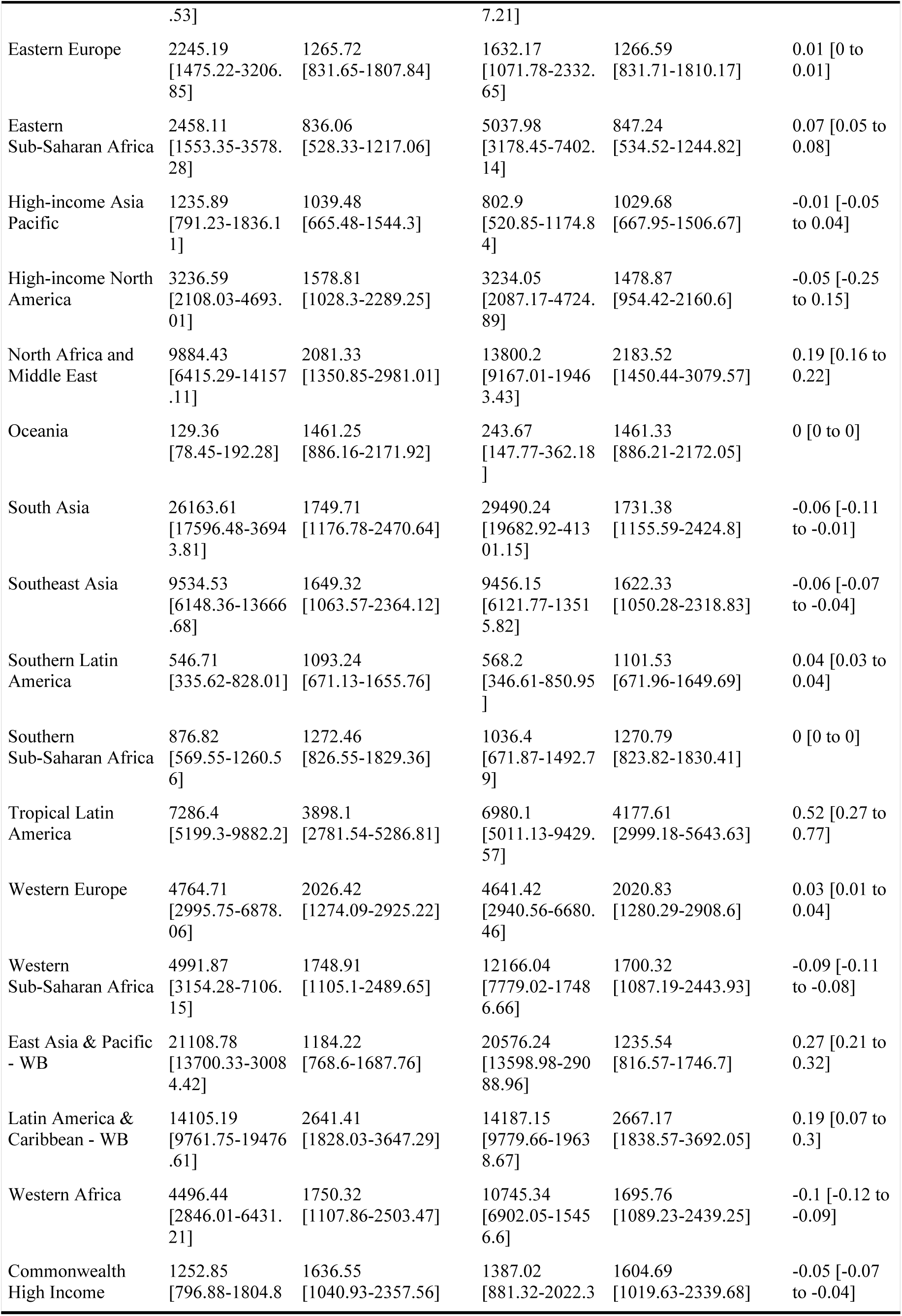
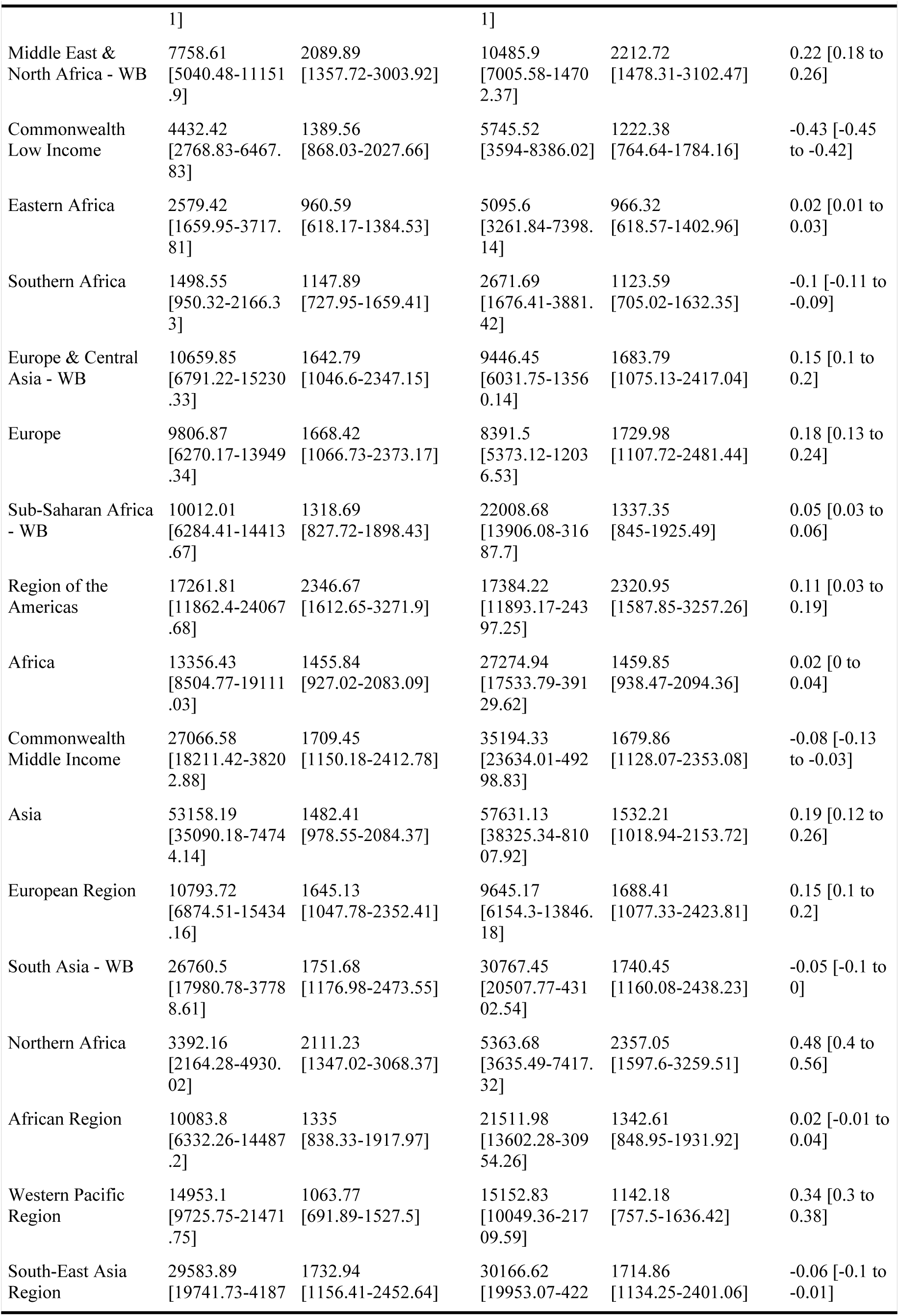
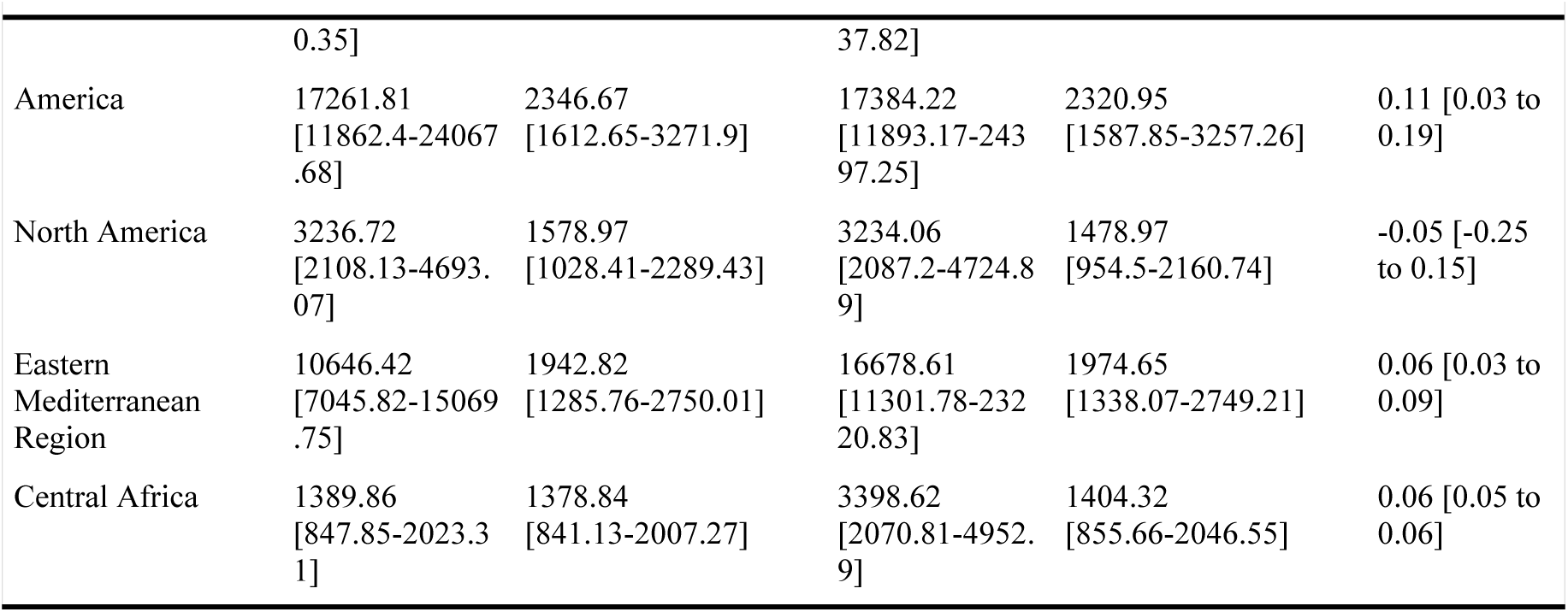
Global incidence, age-standardized incidence rate (ASIR), and estimated annual percentage changes (EAPCs) of ASIR of migraine from 1990-2021.

Based on the comprehensive global data from 1990 to 2021, the ASIR of ADHD exhibited a slight but consistent downward trend, declining from 404.4 to 386.7 per 100,000 population (**Table 2**). This overall decrease was reflected in an EAPC of -0.36% (−0.43 to -0.29). In terms of absolute disease burden, the number of incidence globally showed a contrasting increase due to population growth, rising from approximately 2,360,027 in 1990 to 2,657,117 in 2021. A clear sex disparity persisted throughout the period. While the ASIR decreased for both females (EAPC = -0.34%) and males (EAPC = -0.37%), the ASIR in males remained more than double that of females (573.1 vs. 226.3 per 100,000 in 1990; 547.8 vs. 215.0 per 100,000 in 2021).

**Table 2.**
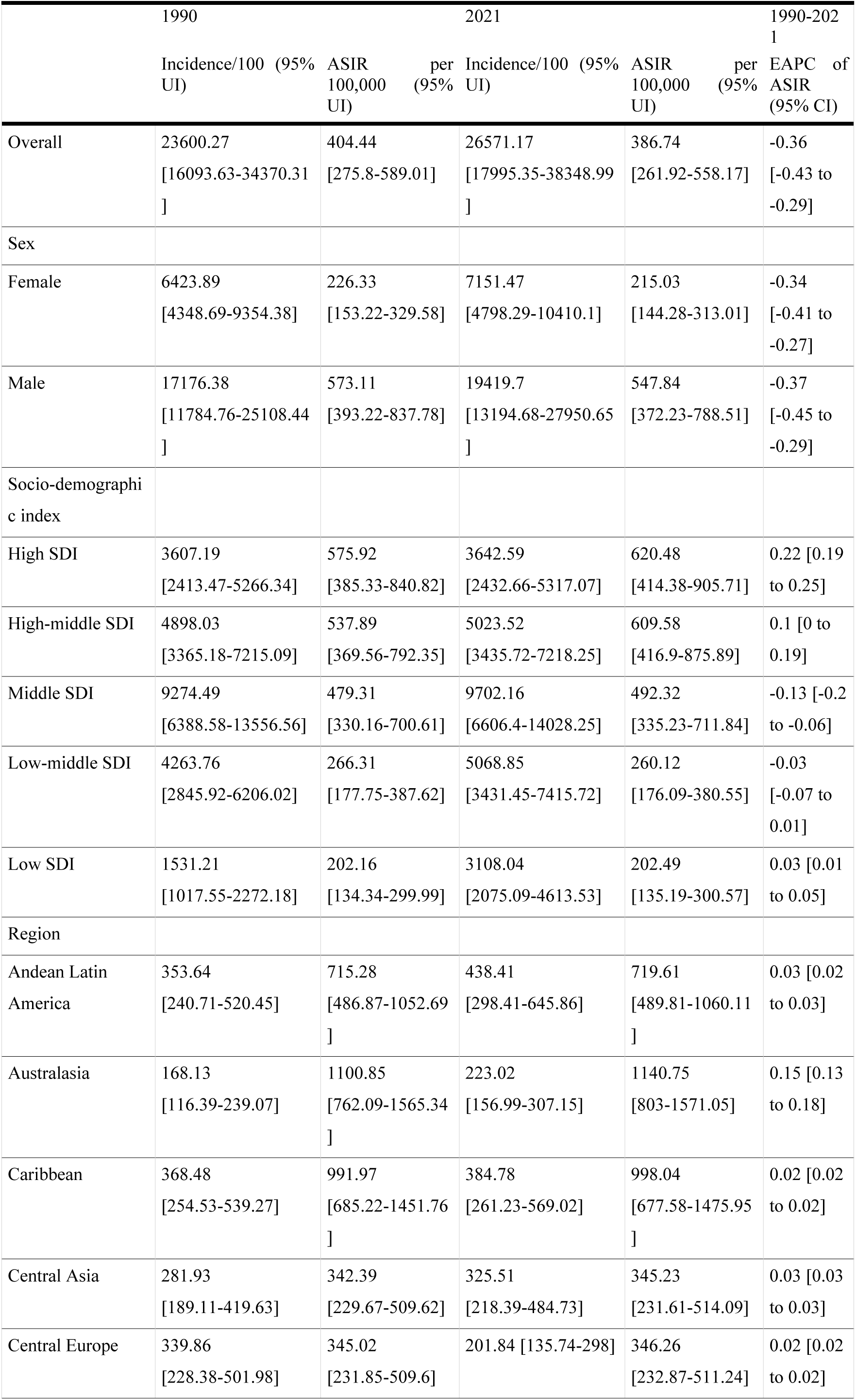
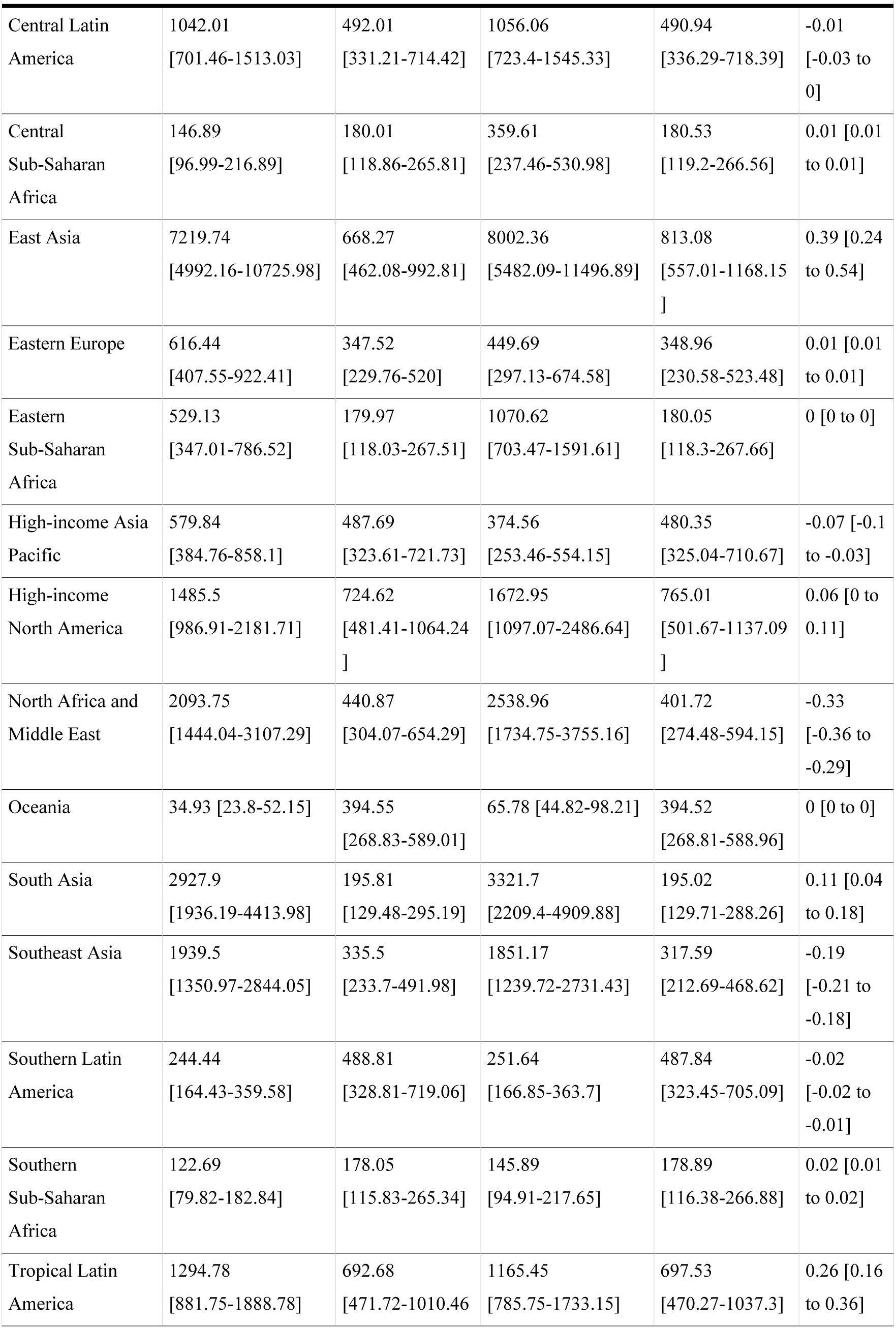
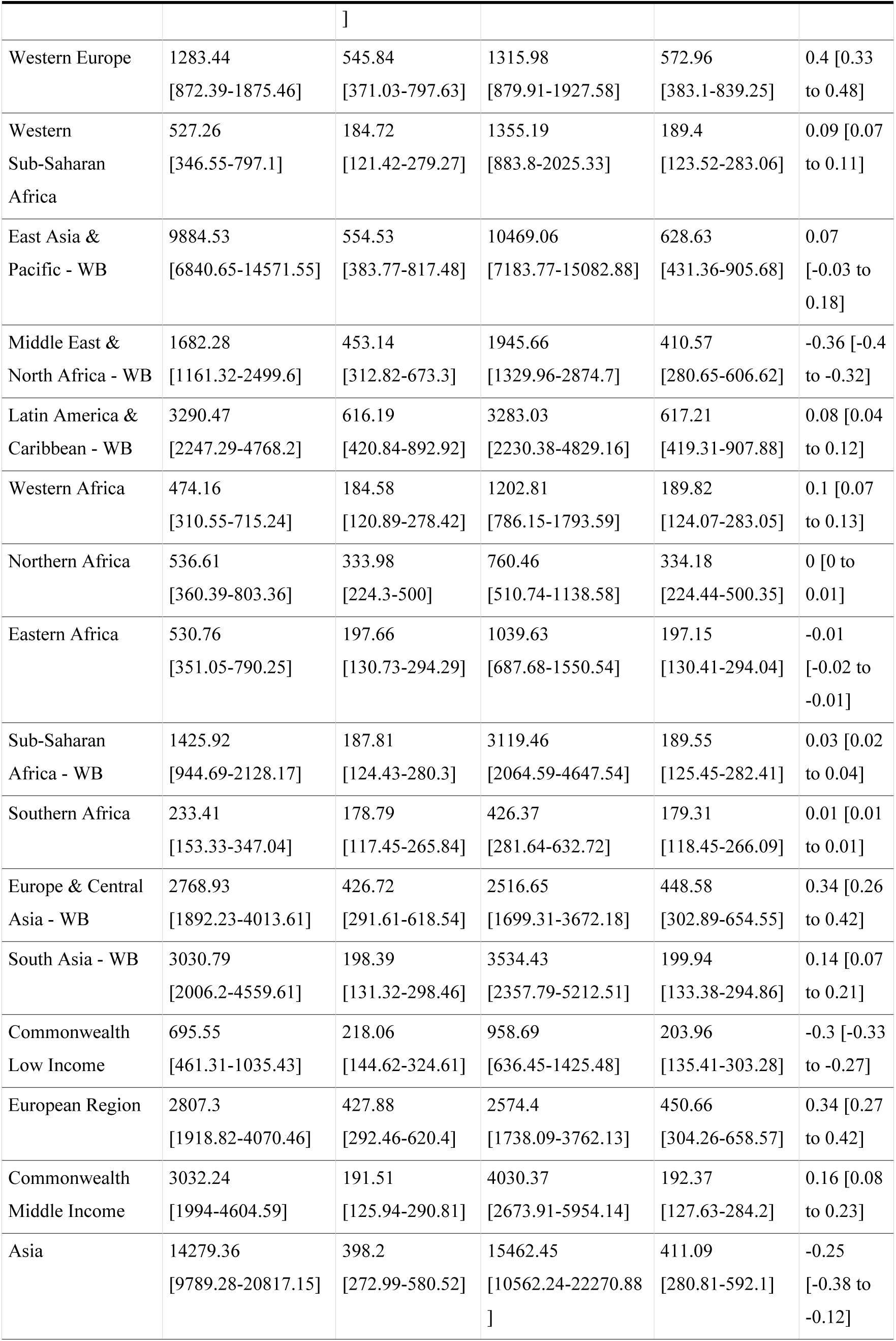
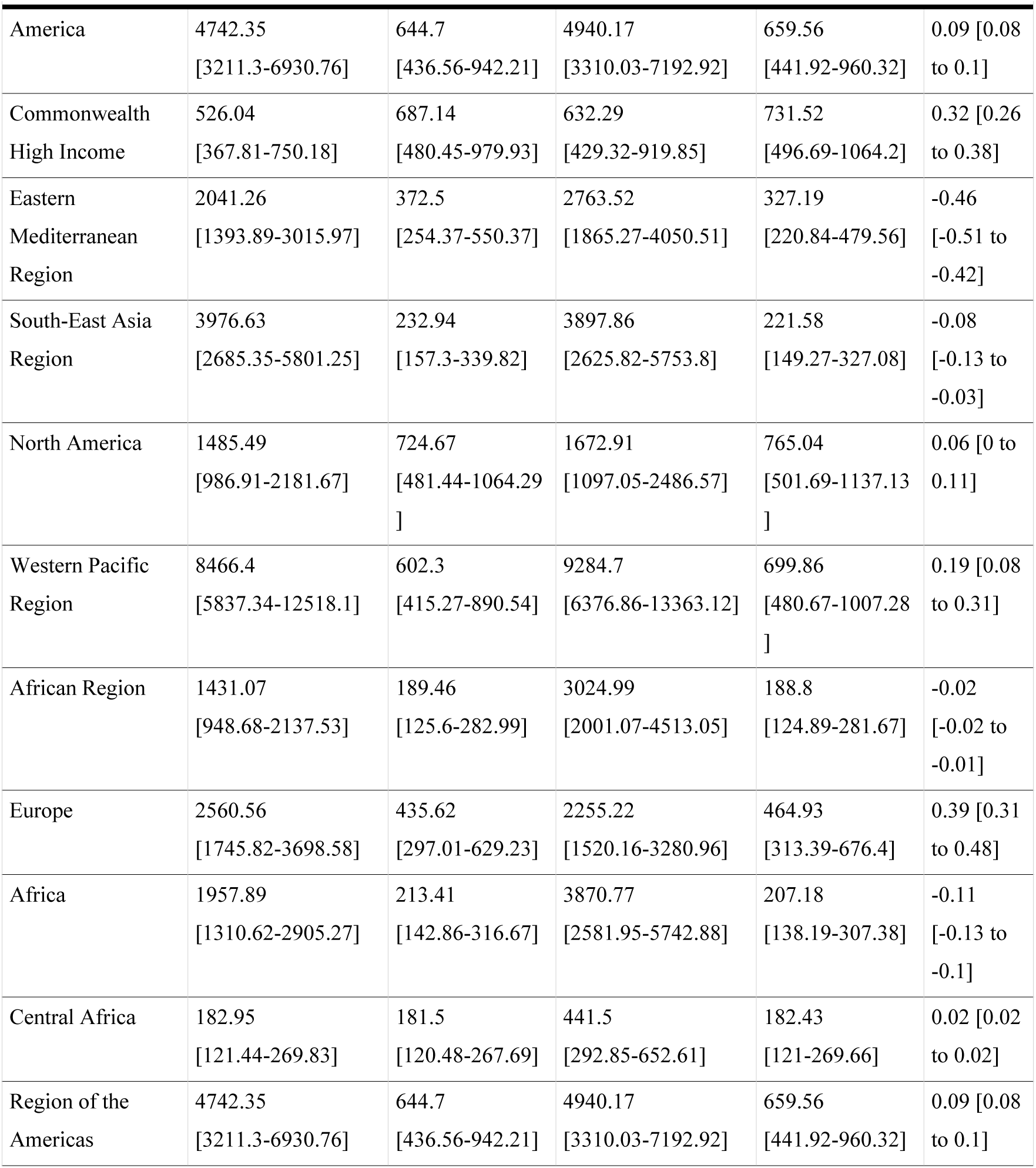
Global incidence, age-standardized incidence rate (ASIR), and estimated annual percentage changes (EAPCs) of ASIR of attention deficit hyperactivity disorder (ADHD) from 1990-2021.

When stratified by the SDI, ADHD incidence trends revealed a clear divergence correlated with development levels: high-SDI regions experienced a notable increase in ASIR (EAPC = 0.22%), a trend also observed though more modestly in high-middle SDI regions (EAPC = 0.10%). In contrast, middle-SDI regions exhibited a clear decline (EAPC = -0.13%), while low-middle SDI regions remained relatively stable (EAPC = -0.03%). Meanwhile, low-SDI regions registered a mild increase (EAPC = 0.03%). This pattern suggests a shift in the epidemiological landscape of ADHD, wherein reported incidence—and thus apparent disease burden—is rising in highly developed regions, likely due to enhanced awareness and diagnostic practices, while declining or stabilizing in many developing regions.

### 3.2 Co-occurrence patterns of migraine and ADHD

Based on co-occurrence patterns of migraine and ADHD, in 2021, countries and regions were categorized into three distinct groups: the majority exhibited an ADHD-dominant pattern, predominantly located in high-SDI regions; a consistent pattern was mainly observed across South Africa and the Middle East, where both diseases showed comparable prevalence levels, while a migraine-dominant pattern was in North Africa (**Figure 2A**). Cross-tabulation analysis indicated that the burden of both diseases remained generally consistent from 1990 to 2021 with in 2021, though a gradual shift toward the ADHD-dominant pattern has been observed in recent years (**Figure 2B-C**). Overall, a positive correlation was observed between the incidence of migraine and ADHD (*P* < 0.001). This correlation was strongest in the consistent regions (r = 0.894), moderate in the migraine-dominant regions (r = 0.701), and weak to moderate in the ADHD-dominant regions (r = 0.40) (**Figure 3**). Over time (1990–2021), most country-year points shifted from the lower-left to the upper-right quadrant of the scatter plot, indicating a synchronous increase in the burden of both diseases. ADHD-dominant regions demonstrated a higher ADHD burden at comparable levels of migraine, while the migraine-dominant regions showed the opposite pattern. These trajectory patterns reflect heterogeneous growth in comorbid profiles across regions and over time, with the consistent group exhibiting the most stable and strongest co-varying trend.

**Figure 2.**
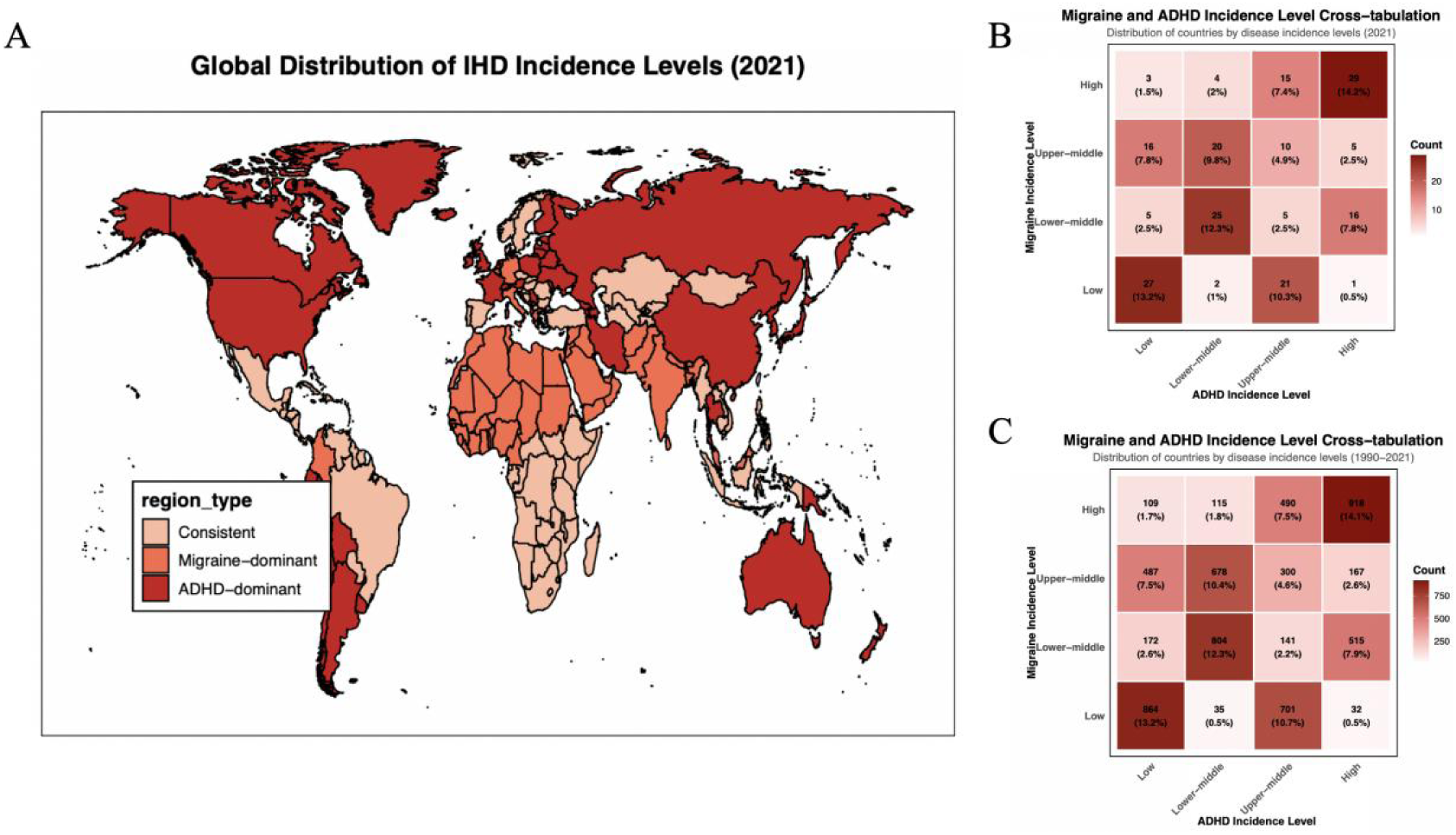
Co-occurrence patterns of migraine and ADHD. (**A**) Global distribution of co-occurrence patterns of migraine and ADHD. (**B**) Cross-tabulation analysis of migraine and ADHD incidence in 2021. (**C**) Cross-tabulation analysis of migraine and ADHD incidence from 1990 to 2021.

**Figure 3.**
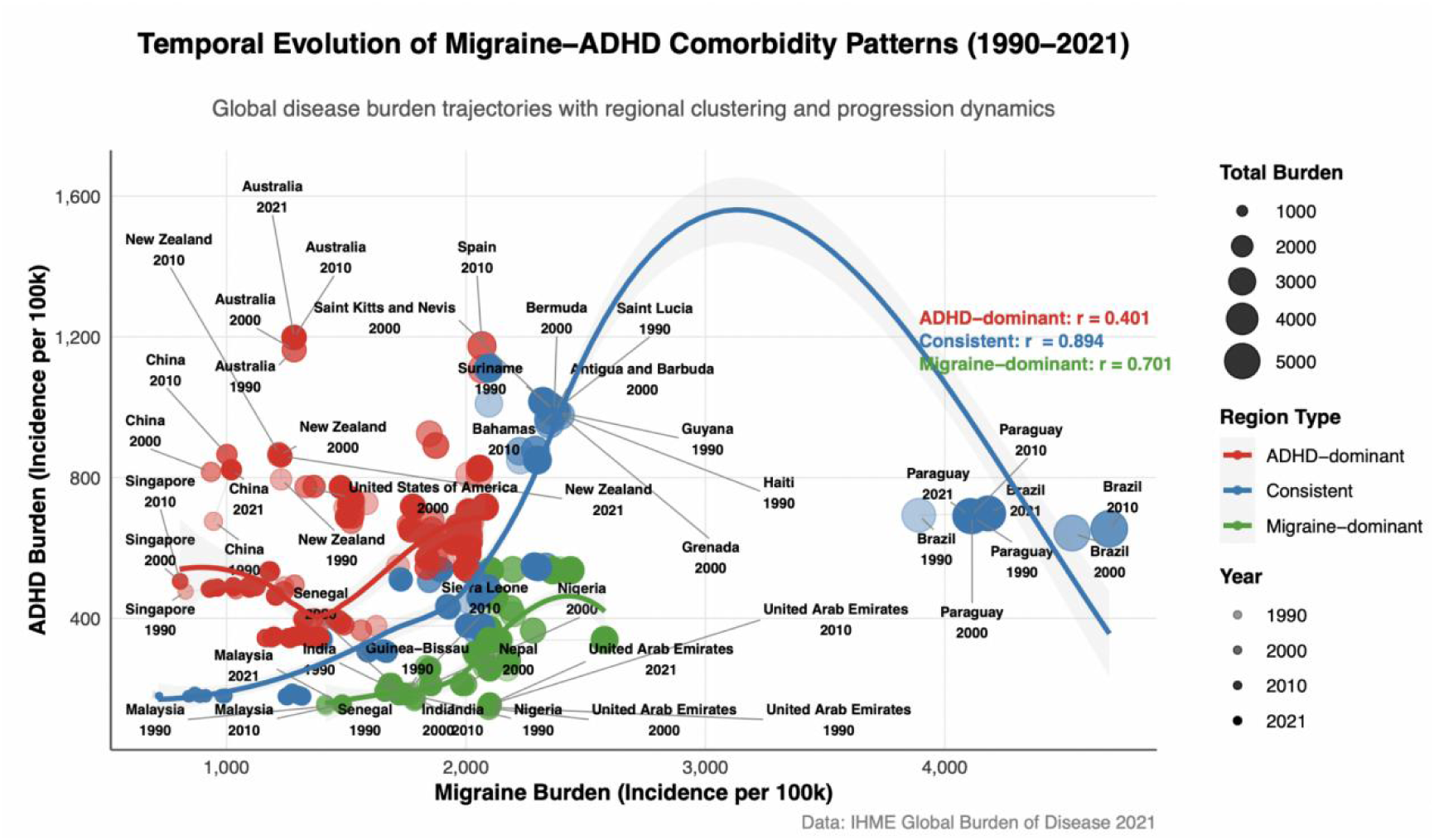
Correlation analysis between migraine and ADHD incidence.

### 3.3 Co-occurrence patterns in global groupings by economy

In developed countries, the co-occurrence disease burden was predominantly moderate to severe and has shown a general trend of alleviation over time, though exceptions exist—such as Spain, which reported relatively critical in earlier years. In emerging economies, the burden largely remained at moderate to severe levels. Other developing countries generally sustained a moderate to severe burden, with signs of further escalation in certain regions during recent periods. Least developed countries consistently exhibited mild to moderate burden levels with relative stability over time (**Figure 4**). The patterns of co-occurrence in some countries showed obvious similarities, including Haiti, Grenada, Bahamas, and Bermuda (**Figure 5**). Overall, these patterns indicate a clear correlation between the severity of co-occurrence patterns of migraine and ADHD and levels of economic development, alongside notable regional and temporal heterogeneity.

**Figure 4.**
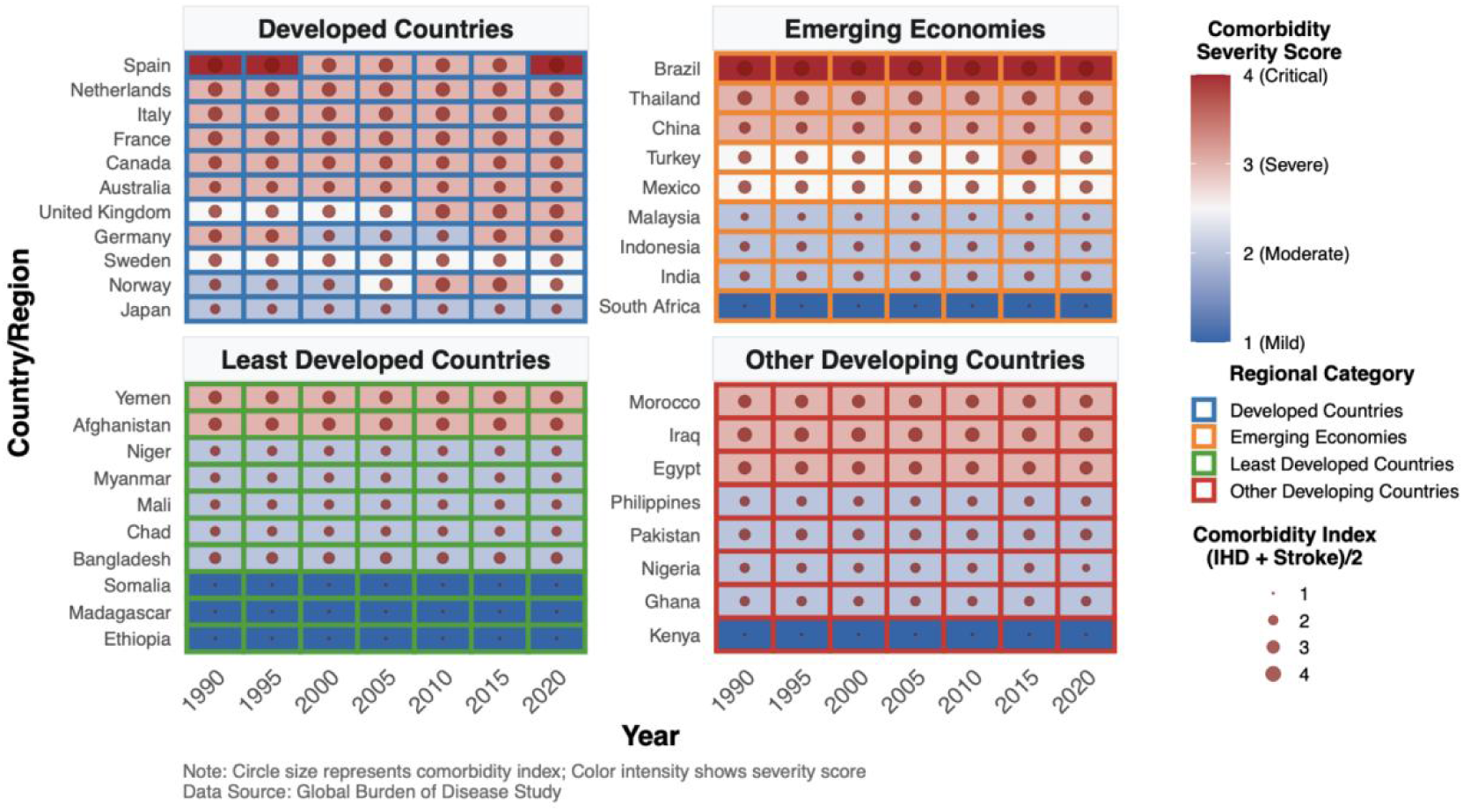
Co-occurrence patterns of migraine and ADHD in countries classified by development stage.

**Figure 5.**
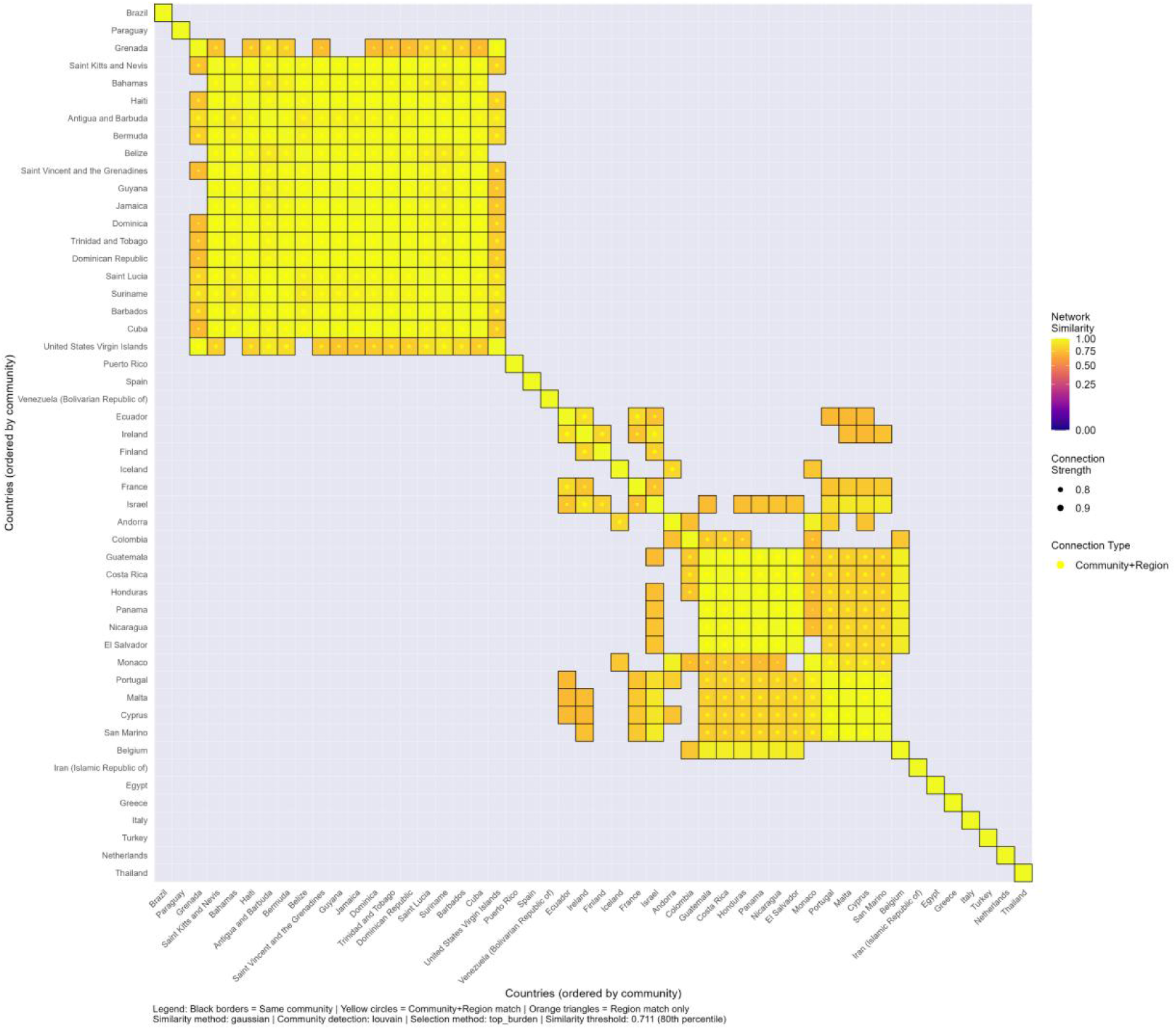
Country similarity network based on migraine and ADHD incidence.

### 3.4 Identification of risk factors under co-occurrence patterns and their spatial distribution

Among the three co-occurrence pattern groups, elevated temperature exposure, iron deficiency, and metabolic risks consistently stood out as major contributing factors. Each of these factors showed positive SHAP values, suggesting that higher exposure levels corresponded to increased risk. Behavioral risks, such as high body-mass index (BMI) and tobacco consumption, played a significant role in consistent and ADHD−dominant groups. In contrast, ambient particulate matter pollution and non-optimal temperature conditions had a more pronounced impact on migraine−dominant group (**Figure 6**). Geographically, regions spanning North Africa, the Middle East, and South Asia experienced the most notable effects of high and non-optimal temperatures. Iron deficiency was predominantly concentrated in Sub-Saharan Africa. Metabolic risks, including high BMI and tobacco use, were more prevalent throughout Europe, the Americas, and certain areas of East Asia. Meanwhile, exposure to ambient particulate pollution was most significant in South and East Asia, as well as parts of Africa. The nine most influential risk factors, consistently ranked across all groups, were chosen for representation in global choropleth maps (**Figure 7**). These spatial distributions highlight shared environmental and metabolic influences across all co-occurrence groups, while behavioral and pollution-related risks reveal more distinct, group-specific, and geographically varied patterns.

**Figure 6.**
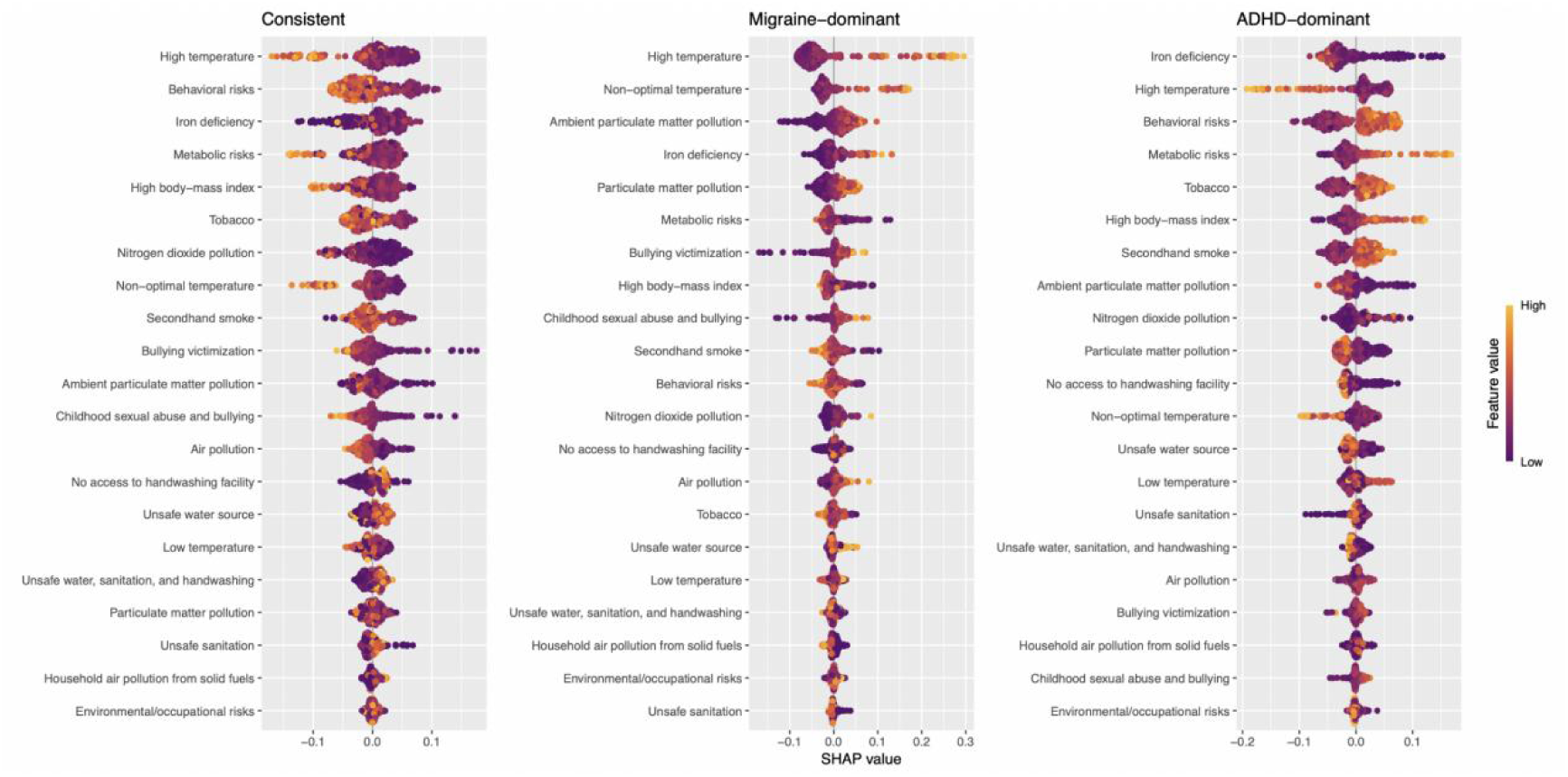
Risk factor identification under co-occurrence patterns of migraine and ADHD.

**Figure 7.**
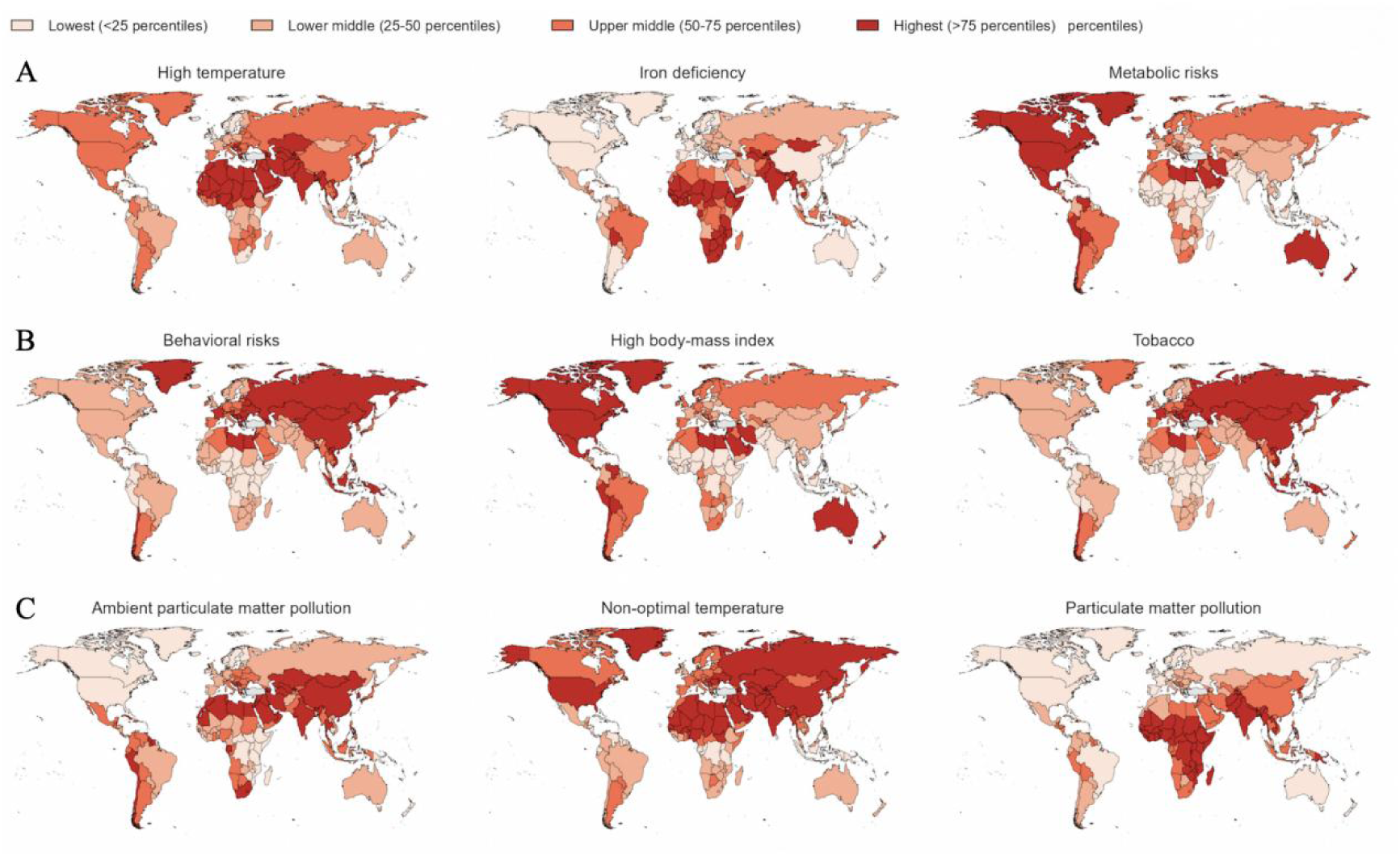
Global spatial distribution of major risk factors associated with migraine and ADHD co-occurrence patterns.

### 3.5 Future forecasts of global burden of migraine and ADHD

Historically, the ASIR of migraine was significantly higher in females than in males. The ASIR exhibited a pattern of initial increase followed by a subsequent decline, reaching its peak around the year 2010 across both sexes. ARIMA model-based central projections suggested that the overall ASIR will remain approximately stable through 2050, with the gender gap persisting. Uncertainty increases over time, as indicated by progressively widening PIs (**Figure 8A-B**). The ASIR of ADHD was consistently higher in males than in females (**Figure 8C-D**). Between 1990 and 2021, male ASIR initially increased and then declined, stabilizing after 2005, while female rates showed minor fluctuations. Throughout the forecast period (2022–2050), both sexes are projected to experience stable or mildly increasing trends. Although the median forecast suggested limited overall change, substantial uncertainty is anticipated in the long term.

**Figure 8.**
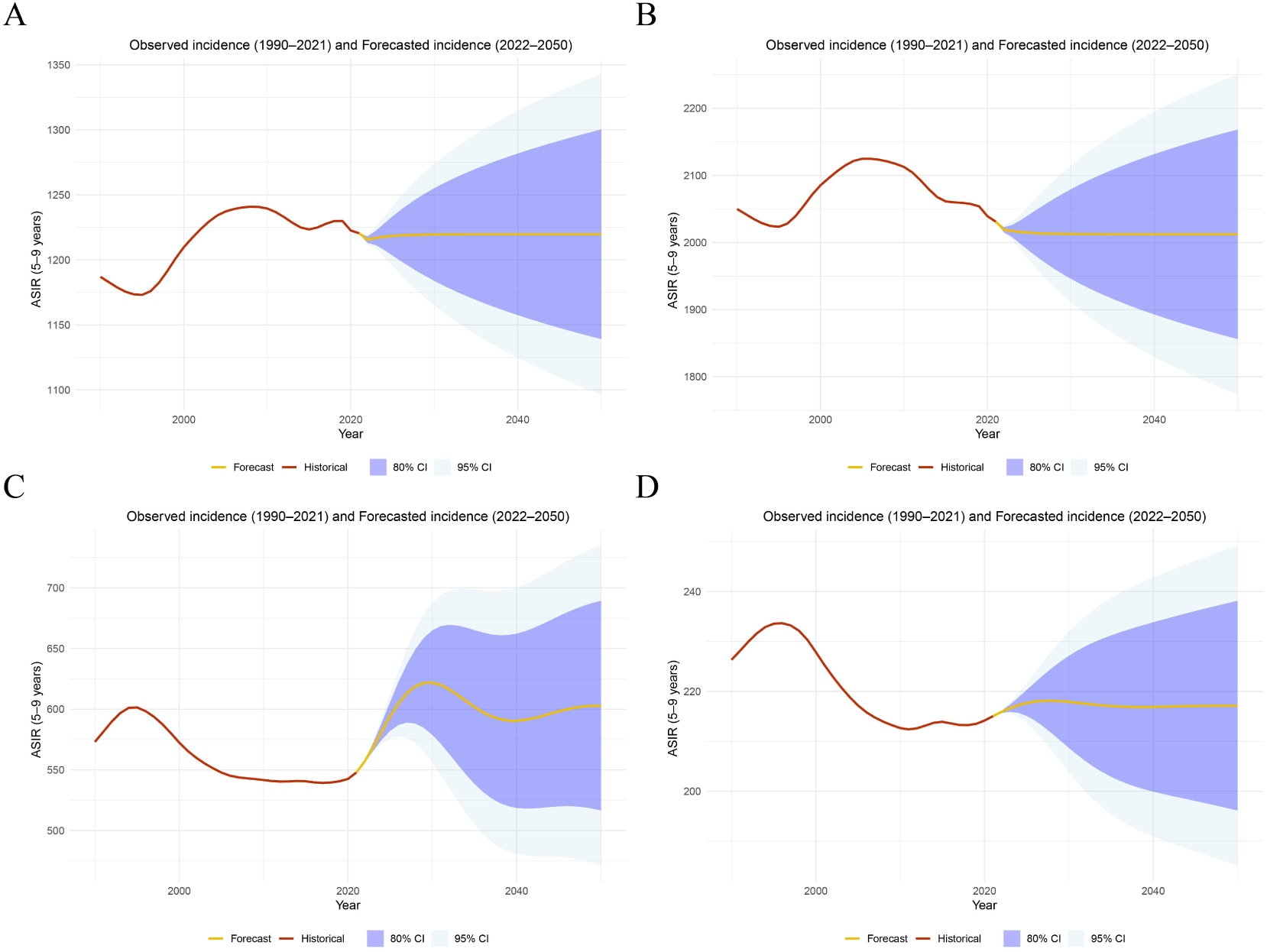
Future forecasts of global burden of migraine and ADHD. (**A**) ARIMA model-based central projections for male migraine. (**B**) ARIMA model-based central projections for female migraine. (**C**) ARIMA model-based central projections for male ADHD. (**D**) ARIMA model-based central projections for female ADHD.

## 4 Discussion

Based on GBD 2021, we delineated three reproducible ADHD–migraine co-occurrence patterns: ADHD-dominant (enriched in high-SDI settings), consistent pattern (concentrated in southern Africa and the Middle East), and migraine-dominant (represented by North Africa), all showing positive covariance over 1990–2021. The level of SDI is closely related to disease detection ability, accessibility to diagnosis and treatment, and health literacy, which in turn shapes different comorbidity structures: High SDI areas are more likely to identify neurodevelopmental disorders such as ADHD, while low and medium SDI areas are exposed to higher migraine burden due to environmental and nutritional factors (such as high temperature, iron deficiency). These interpretations align with the SDI framework and recent GBD syntheses that document development-linked heterogeneity in neurologic burdens and risk attribution ^[5][15]^.

Regrading risk factors, environmental exposure, iron metabolism and behavioral metabolic pathways are involved in the co-morbidity mechanism of the two diseases. High temperature not only reduces the threshold of migraine attack by inducing dehydration, interfering with sleep and vascular reactivity, but also indirectly affects ADHD symptoms due to increased cognitive load, suggesting the necessity of climate adaptive intervention ^[16][17][18]^. Iron is a convergent modulator of catecholamine synthesis and myelination. Meta-analytic and imaging evidence indicates reduced ferritin and lower brain iron in pediatric ADHD, with signals of symptom improvement in iron-repletion studies^[19]^. In migraine, iron biology appears bidirectional—systemic deficiency is associated with susceptibility, whereas regional brain iron accumulation has been linked to chronification, suggesting that perturbed iron homeostasis can sensitize pain networks and executive circuits in parallel^[20]^. In addition, metabolic behavior risk factors such as obesity and tobacco play a role in both diseases: obesity exacerbates migraine through low-grade inflammation, CGRP upregulation, and sleep fragmentation, while there is a two-way promotion relationship between ADHD and obesity; smoking during pregnancy is not only associated with increased risk of migraine, but also closely related to the occurrence of ADHD in offspring ^[21][22][23][24][25]^.

The regional distribution of comorbidity aligns with pollution patterns, as higher PM exposure in East Asia, South Asia, and parts of Africa has been linked to increased migraine-related emergency visits and elevated ADHD risk in systematic reviews ^[26][27][28]^. Mechanistically, pollution may exacerbate pain and attentional pathways through oxidative stress, neuroinflammation, and blood–brain barrier disruption, with combined heat and pollution further lowering activation thresholds ^[29]^. At the level of neurobiology, both types of diseases have dysfunction of monoaminergic system, but the specific mechanisms are different. Migraine is centered on CGRP-glutamic acid-trigeminal neurovascular pathway, and the effectiveness of its targeted drugs verifies the causal role of this pathway^[30][31]^. ADHD is mainly associated with dopamine system dysfunction and is supported by genetic evidence of transporters and receptors ^[32][33]^. Region-specific modifiers such as iron deficiency, heat stress, and pollution-induced inflammation/oxidative stress may amplify glutamatergic excitotoxicity and dopaminergic vulnerability, thereby converging to align the epidemiologic trajectories of the two disorders in consistent regions. Notably, distinct patterns also reveal specific triggers: migraine-dominant regions are more strongly influenced by non-optimal temperature and particulate matter, underscoring the primacy of environmental drivers of acute attacks, whereas ADHD-dominant regions reflect the combined impact of metabolic and behavioral risks such as high BMI and tobacco use together with higher ascertainment. This ’common-specific’ framework suggests that public health strategies should be precisely designed in combination with geography and risk pedigrees, such as prioritizing thermal adaptation and air quality management in hot and arid areas and strengthening weight control and tobacco prevention and control in high SDI urban environments.

Based on GBD 2021 data, this study systematically evaluated the disease burden and key risk factors of migraine and ADHD comorbidity, and predicted its development trend to 2050. However, our study still has some limitations. First, GBD research itself depends on the quality and unified coding of the original data, and there may be a risk of insufficient identification in low SDI areas. Second, the ecological exposure indicators used are difficult to capture the micro-environmental differences, and the statistical correlation is susceptible to residual confounding factors, so it is impossible to directly infer causality. In addition, the predictive analysis itself is accompanied by the accumulation of uncertainty, and structural shocks (such as pandemics, changes in diagnostic criteria, extreme high temperatures, or wildfire events) may deviate from the standard trajectory based on EAPC/time series. Final, although we identified iron metabolism and other metabolic pathways as potential common levers for the regulation of comorbidity, the specific mechanism remains to be further verified by longitudinal imaging and intervention trials.

## 5 Conclusion

ADHD and migraine worldwide show a regular pattern of comorbidity related to the level of regional development. Common risk factors such as high temperature exposure, iron deficiency, and metabolic/behavioral risks may reduce the incidence thresholds of the two diseases, while specific environmental exposure (such as particulate matter pollution) and health system factors affect their dominant types. The above findings support the implementation of a comprehensive public health strategy, covering climate adaptation, nutrition improvement, metabolic health management, tobacco control and diagnostic specification optimization.

## Data availability

The data used in this study were obtained from the Global Burden of Disease Study 2021 (GBD 2021), which is publicly available through the Global Health Data Exchange (GHDx) and the GBD Results Tool provided by the Institute for Health Metrics and Evaluation (IHME) (https://ghdx.healthdata.org/gbd-2021).

## Author contributions

XW conceived the study, performed the data collection and bioinformatics analysis, interpreted the results, and drafted the manuscript. JJ assisted with the study design and contributed to manuscript revision. All authors have read and approved the final manuscript.

## Data Availability

The data used in this study were obtained from the Global Burden of Disease Study 2021 (GBD 2021), which is publicly available through the Global Health Data Exchange (GHDx) and the GBD Results Tool provided by the Institute for Health Metrics and Evaluation (IHME).

https://ghdx.healthdata.org/gbd-2021

## Acknowledgements

Not applicable.

